# Associations of autism diagnosis, traits, and genetic liability with subsequent night-time sleep duration trajectories from infancy to adolescence

**DOI:** 10.64898/2026.03.10.26348028

**Authors:** Reesha Zahir, Sarah Moody, Isabel Morales-Muñoz, Aja Louise Murray, Sue Fletcher-Watson, Alex S.F. Kwong, Daniel J. Smith

**Author notes:** Corresponding authors: Reesha Zahir -, Alex S.F. Kwong -, Address: Division of Psychiatry, University of Edinburgh, Chancellor’s Building, 49 Little France Crescent, Edinburgh, EH16 4SB. Denotes shared authorship (Reesha Zahir, Sarah Moody, Sue Fletcher-Watson, Alex S.F. Kwong, Daniel J. Smith). **Author Contributions** Conceptualisation: R.Z., S.F.W., A.S.F.K, and D.J.S. Methodology and formal analysis: R.Z., A.S.F.K., I.M.M., and A.L.M. Supervision: S.F.W., A.S.F.K, and D.J.S. Funding acquisition: R.Z. Writing – original draft: R.Z. and S.M. Writing – review and editing: all authors. **Acknowledgements** We are grateful to all the families who took part in this study, the midwives for their help in recruiting them, and the whole ALSPAC team, which includes data collection staff, data and administrations staff, technical managers and the technical staff with the Bristol Bioresource Laboratory, based within the University of Bristol. **Conflicts of interest** The authors report no conflicts of interest. **Ethical Approval** Ethical approval for the study was obtained from the ALSPAC Law and Ethics committee and local research ethics committees (NHS Haydock REC: 10/H1010/70).

## Abstract

**Background:** Autistic individuals experience higher rates of sleep problems throughout their lives, and there is considerable heterogeneity in manifestations of these issues that remains unexplained. Here, we examine associations over time of heterogenous sleep trajectories with autism diagnosis, and behavioural and genetic factors related to autism.

**Method:** We used data from the Avon Longitudinal Study of Parents and Children (N=13,886, autistic n=150). The primary outcome was parent and self-reported night-time sleep duration, measured on 10 occasions (between 0.5y and 15.5y). The independent variables were autism diagnosis, autism polygenic score (PGS) and four parent-reported autistic traits: repetitive behaviour, social communication, speech coherence, and sociability. Latent class growth analysis was conducted to identify heterogenous classes of sleep trajectories, and these trajectory classes were regressed onto the independent variables.

**Results:** Four night-time sleep duration trajectory subclasses were identified; shorter (n=512, 4.1%), longer (n=1654, 13.1%), intermediate-shorter (n=3630, 28.8%), and intermediate-longer (used as the reference class; n=6825, 54.1%). An autism diagnosis was associated with a shorter or intermediate-shorter sleep duration trajectory, compared to the reference class. Similarly, higher scores in domains of repetitive behaviour, speech coherence and social communication were associated with shorter sleep duration trajectories. The autism PGS and sociability were not associated with any sleep trajectories compared to the intermediate-longer sleep trajectory (reference group).

**Conclusion:** An autism diagnosis and specific autistic traits were associated with poorer long-term sleep outcomes across childhood and adolescence, highlighting the need for early, sustained sleep interventions, and the potential of trait-specific mechanisms for sleep problems.

**Highlights:**

- Four distinct night-time sleep duration trajectories were identified across development
- Autism diagnosis predicted shorter and intermediate-shorter sleep trajectories
- Specific (but not all) autistic traits were linked to shorter sleep trajectories
- Autism PGS did not predict sleep duration trajectories

## Introduction

Autism is a heterogenous lifelong neurodevelopmental condition characterised by distinctive profiles of social communication, repetitive behaviour, and sensory domains (American Psychiatric Association, 2013). Sleep problems are highly prevalent among autistic individuals, affecting an estimated 40-83% of autistic children (Richdale & Schreck, 2009; Schwichtenberg et al., 2022), with similar difficulties also observed in autistic adolescents and adults (Díaz-Román et al., 2018; Lugo et al., 2020). Sleep difficulties in this population are associated with substantial mental and physical burden, including poorer mental health, adverse physical health outcomes, and poorer developmental trajectories across domains such as language, social functioning, sensory processing, and motor skills (Foster et al., 2023; Henderson et al., 2023; Lawson et al., 2020; Levin et al., 2022; Tudor et al., 2012; Veatch et al., 2017). Meta-analyses indicate that autistic children experience a broad range of sleep problems, yet there is substantial heterogeneity in both the type and persistence of these difficulties across development (Elrod & Hood, 2015). Despite this, most research has focused on cross-sectional average group differences, providing limited insight into individual variation in long-term sleep patterns.

Given that autism is understood as a neurodevelopmental condition with multiple underlying factors (Chaste & Leboyer, 2012), it is plausible that distinct mechanisms may also underpin co-occurring sleep problems, leading to heterogeneous manifestations (Schreck & Richdale, 2020). Although several behavioural and biological risk factors for sleep problems have been proposed (Hu et al., 2009; Mazzone et al., 2018; Schreck & Richdale, 2020), the mechanisms underpinning sleep difficulties in autistic individuals remain poorly understood. Longitudinal approaches that capture change over time may be particularly informative for a fuller understanding of sleep as a dynamic process that evolves across childhood and adolescence.

One potential pathway involves shared genetic liability. Recent studies have explored associations between autism between autism polygenic scores (PGS) and sleep, based on the hypothesis that genes associated with autism may also be enriched for variation affecting sleep (Grove et al., 2019). However, findings have been mixed. Two studies have reported no significant associations between autism PGS and sleep disturbances (Niarchou et al., 2022; Ohi et al., 2021), whereas a more recent study identified an association between autism PGS and evening chronotype (Fahey & Lopez, 2024). No studies to date have examined whether an autism PGS is associated with long-term sleep trajectories, which may provide greater insight into developmental patterns of sleep than cross-sectional outcomes.

Behavioural pathways have also been hypothesised, with different autistic traits proposed to have specific contributions to the development and persistence of sleep difficulties (Elkhatib Smidt et al., 2022; Mazzone et al., 2018). For example, restrictive and repetitive behaviours may increase distress around changes to bedtime routines, delaying sleep onset (Reynolds & Malow, 2011), and social difficulties may reduce participation in activities that support circadian regulation e.g., sports (Geoffray et al., 2016). Communication difficulties may also complicate bedtime routines by limiting understanding of parental instructions (Ibañez et al., 2018; Mazzone et al., 2018). Interestingly, a recent cross-sectional study identified two distinct groups of children based on their autistic traits and sleep problems: a younger group that had insufficient sleep, sensory sensitivities, and hyperactive behaviour, and an older group that experienced excessive daytime sleepiness, increased cognitive issues, and mood symptoms - suggesting that specific autistic traits may have different influences on sleep outcomes, which may also change over time (Lenker et al., 2025). Although there is evidence that sleep problems change across the lifespan in autistic individuals (Baker et al., 2023; Humphreys et al., 2014; Jovevska et al., 2020; Verhoeff et al., 2018), most studies linking autistic traits to sleep problems are cross-sectional, limiting understanding of how these traits relate to long-term sleep trajectories and whether this varies across development.

Large longitudinal cohorts provide a valuable opportunity to address these gaps by identifying heterogeneous sleep trajectories and examining factors that predict membership in different developmental patterns (Nagin & Odgers, 2010). To date, only one large-scale cohort study has examined heterogeneity in sleep trajectories in relation to autism, finding that autism diagnosis and higher levels of autistic traits were associated with an increased likelihood of having trajectories characterised by increasing sleep disturbance over this period. However, this study focused on a narrow developmental window in early childhood (1.5 to 9 years), and examined the relationship with overall autistic trait level, without discriminating between specific traits.

We aimed to identify distinct night-time sleep duration trajectories from infancy through adolescence in the Avon Longitudinal Study of Parents and Children (ALSPAC), a UK-based general population cohort. Previous studies have identified heterogeneity in night-time sleep duration trajectories in ALSPAC (Manitsa et al., 2024; Morales-Muñoz et al., 2024). Building on this work, we examined the relationship between autism and heterogenous sleep trajectories, which has not previously been explored over such an extended period across development. The primary aim of our study was to identify subclasses of night-time sleep duration trajectories and examine whether autism diagnosis was associated with specific sleep trajectory subclasses. We hypothesised that an autism diagnosis would be associated with shorter sleep duration trajectories relative to the average. Secondary aims were to examine whether autism PGS, as well as four key autistic traits – social communication, speech coherence, repetitive behaviour, and sociability – were associated with sleep trajectory subclasses, to better understand how genetic and behavioural factors related to autism may contribute to long-term sleep patterns. Since these factors can be measured across the general population, they offer a valuable opportunity to identify associations between specific elements of autistic profiles and sleep problems. Given the limited and inconsistent existing literature, no specific hypotheses were made regarding individual trait and autism PGS associations.

## Methods

### Study Cohort

The data used in the current study is from the ALSPAC, a longitudinal birth cohort based in Avon, a region in the South-West of England (Boyd et al., 2013; Fraser et al., 2013). Pregnant women in the region with expected dates of delivery between 1^st^ April 1991 and 31^st^ December 1992 were invited to take part. There was a total of 14,676 foetuses, resulting in 14,062 live births and 13,988 children who were alive at 1 year of age. When the oldest children were approximately 7 years, additional participants were recruited to bolster the sample, resulting in 15,447 pregnancies, 15,658 lives births, and 14,901 children alive one year later. At age 18, study children were sent’fair processing’ materials describing ALSPAC’s intended use of their health and administrative records and were given clear means to consent or object via a written form. Data were not extracted for participants who objected, or who were not sent fair processing materials.

Further information about the ALSPAC study can be found at their website (https://www.bristol.ac.uk/alspac/) and complete information about the study variables collected in the study are available on the ALSPAC website (http://www.bristol.ac.uk/alspac/researchers/our-data/). Ethical approval was obtained from the ALSPAC Ethics and Law Committee and the Local Research Ethics Committees (NHS Haydock REC: 10/H1010/70), and informed consent for the use of data was obtained from participants accordingly.

## Measures

### Ascertainment of Autism Diagnosis

A multi-source approach was used to identify autistic participants in the sample (Pender et al., 2021; Rai et al., 2018; Williams et al., 2004). Participants with an autism diagnosis were identified via any of the following three sources (all other participants in the sample were classified as non-autistic): **i.** A parent-report questionnaire completed at 9.5 years, which asked mothers whether they had even been told that their child had a diagnosis of autism**; ii.** Linked health records (National Health Service) indicating whether participants had received a diagnosis of autism (or related diagnoses) and **iii.** Linked education records (Pupil Level Annual School Census data for England, 2003) indicating whether autism was listed as a primary or secondary concern for those needing special education provisions. Any of these three elements was alone a sufficient criterion for classification as autistic for the purposes of this study.

### Autism Polygenic Score (PGS)

The autism PGS was derived using summary genetic data from a recent genome-wide association study (GWAS) of autism (Grove et al., 2019). Full quality control information on genetic data have been previously reported (Kwong et al., 2021), but are also given in the supplement (see S1). The autism PGS was created using PRSice-2, version 2.1.11 (Choi & O’Reilly, 2019), using clumping and thresholding (clump-kb 250kb, clump-r2 0.1, clump-p 1.0). The PS for p-value thresholds was generated from the original GWAS at 5 × 10−8, 1 × 10−6, 1 × 10−4, 0.001, 0.01, 0.05, 0.1, 0.2, 0.5, and 1. The PGS with a threshold of 1 was used for this analysis based on work showing that it was the most predictive threshold in ALSPAC (Grimes et al., 2024). The autism PGS was standardised to have a mean of 0 and a standard deviation of 1; thus, a higher autism PGS represents higher genetic predisposition to autism.

### Autistic Traits

We focused on the following four traits that were identified as the best predictors of autism diagnosis out of 93 measures related to the main features of autism available in ALSPAC: social communication, speech coherence, repetitive behaviour, and sociability temperament (Steer et al., 2010). Each trait was analysed using both total trait scores and a binary variable identifying participants experiencing the highest traits, defined by a cut-off of approximately 10% (Golding et al., 2017). All traits were parent-reported, and assessment details are described below.

#### Social Communication

Measured at 7.7 years using the Social and Communication Disorders Checklist (SCDC), which measures social reciprocity and verbal/non-verbal characteristics that are frequently associated with autism (Skuse et al., 2005).

#### Speech Coherence

Measured at 9.7 years using the coherence subscale of the parent-report Children’s Communication Checklist (CCC), which measures pragmatic aspects of children’s communication (Bishop, 1998).

#### Repetitive Behaviour

Measured at 5.8 years using a four-item ALSPAC questionnaire that asked parents about their child’s repetitive behavioural patterns e.g., repeatedly rocking head or body, having a tic or twitch, etc.

#### Sociability Temperament

Measured at 3.2 years using the sociability subscale of the Emotionality, Activity and Sociability (EAS) scale (Buss & Plomin, 1984), which measures a preference of the child to be in the presence of others to being alone.

### Night-time Sleep Duration

Night-time sleep duration was derived from bedtimes and waking times reported on 10 occasions. These were parent-reported at the first nine assessments (from 6 months to 12 years), and self-reported at the final assessment (15.5 years). See Table 1 for further detail on these measurements.

**Table 1.**
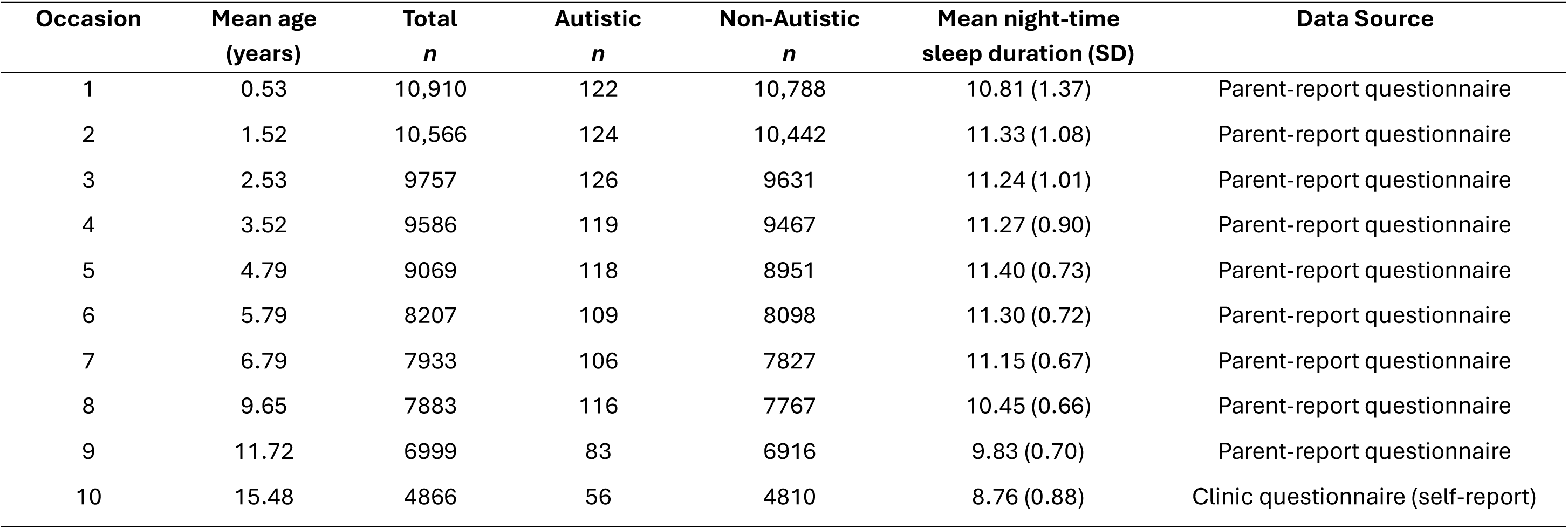
Descriptive statistics of night-time sleep duration.

| **Occasion** | **Mean age**<br>**(years)** | **Total**<br>***n*** | **Autistic**<br>***n*** | **Non-Autistic**<br>***n*** | **Mean night-time**<br>**sleep duration (SD)** | **Data Source** |
| --- | --- | --- | --- | --- | --- | --- |
| 1 | 0.53 | 10,910 | 122 | 10,788 | 10.81 (1.37) | Parent-report questionnaire |
| 2 | 1.52 | 10,566 | 124 | 10,442 | 11.33 (1.08) | Parent-report questionnaire |
| 3 | 2.53 | 9757 | 126 | 9631 | 11.24 (1.01) | Parent-report questionnaire |
| 4 | 3.52 | 9586 | 119 | 9467 | 11.27 (0.90) | Parent-report questionnaire |
| 5 | 4.79 | 9069 | 118 | 8951 | 11.40 (0.73) | Parent-report questionnaire |
| 6 | 5.79 | 8207 | 109 | 8098 | 11.30 (0.72) | Parent-report questionnaire |
| 7 | 6.79 | 7933 | 106 | 7827 | 11.15 (0.67) | Parent-report questionnaire |
| 8 | 9.65 | 7883 | 116 | 7767 | 10.45 (0.66) | Parent-report questionnaire |
| 9 | 11.72 | 6999 | 83 | 6916 | 9.83 (0.70) | Parent-report questionnaire |
| 10 | 15.48 | 4866 | 56 | 4810 | 8.76 (0.88) | Clinic questionnaire (self-report) |

### Covariates

Detailed information on covariates including sources and measurement occasions can be found in the supplementary materials (Table S1). In brief, the following covariates known to influence autism, as well as childhood and adolescent sleep were included in adjusted analyses: sex, maternal postnatal depression, maternal anxiety, epilepsy diagnosis, and socioeconomic status (assessed via maternal occupational class, highest level of maternal educational attainment, and financial problems from parent-report questionnaires).

## Statistical Analysis

We used latent class growth analysis (LCGA) to identify latent classes of night-time sleep duration trajectories from childhood to adolescence (Jung & Wickrama, 2008). This technique is commonly used to explore heterogeneity in longitudinal growth patterns by identifying distinct subgroups over time (Nagin & Odgers, 2010). Trajectories were modelled using linear terms, with the timings of measurements fixed to the mean sample age on each occasion. We decided to use linear terms based on previous work in ALSPAC using LCGA for night-time sleep duration trajectories (Manitsa et al., 2024), due to convergence issues encountered when attempting to fit the models with higher-order terms (i.e., quadratic terms). Several models were fitted by increasing the number of classes, from two up to five, consistent with recommended practice (Jung & Wickrama, 2008). Statistical criteria included the Bayesian Information Criterion (BIC) and the Vuong–Lo–Mendell–Rubin likelihood ratio test (VLMR). Lower BIC values indicate improved model fit, while a significant VLMR test (*p* ≤ 0.05) suggests that a model with *K* classes provides a better fit than a model with *K*–1 classes. Entropy was used to evaluate classification precision, with values closer to 1 indicating clearer class separation. In addition to statistical fit, models were evaluated for interpretability and clinical plausibility of the identified trajectories, as well as for adequate class sizes, with solutions containing classes representing less than 2% of the sample considered less reliable (Jung & Wickrama, 2008). The final model was selected based on convergence across these criteria rather than any single fit index.

We conducted multinomial logistic regressions to examine associations between the exposures (i.e., autism diagnosis, social communication difficulties, speech coherence, repetitive behaviour, sociability temperament, and autism PGS) and night-time sleep duration trajectories, which was the outcome. For the autistic traits, we examined associations using overall trait scores, as well as a binary variable that identified a’high traits group’ for each trait. We calculated correlations between the autistic traits to examine the degree of overlap and independence among them. Since correlations were observed between traits, separate regressions were conducted to examine associations between trajectory subclasses and each autism factor, resulting in 10 adjusted and unadjusted models. When conducting regressions, the trajectory with the largest class membership was chosen as the reference class because it represented the most normative sleep trajectories, which allowed us to identify predictors of deviation from typical sleep patterns.

All analyses were conducted in MPlus 8. Sample sizes for each model included participants with data available for the factor and at least one measurement of sleep. To account for the differences in sample sizes between models, the analysis was repeated in the sub-sample of participants who had complete case data available for each of the measures (n = 3844). Results are presented in the supplement and were broadly aligned with the main findings (Table S2).

### Missing Data

Full Information Maximum Likelihood (FIML) was used to handle the missing sleep outcome data, and multiple imputation was conducted to handle missing covariate data. For the imputation, 50 imputed datasets were generated, and night-time sleep duration measures, autistic trait measures, autism diagnosis, and autism polygenic score were used as auxiliary variables. Separate models were fit to each imputed dataset, and the results were pooled to obtain overall estimates.

## Results

### Descriptive characteristics

Full demographic information can be found in Table 2. The sample comprised 13,886 non-autistic and 150 autistic participants. There were significantly more male participants (79%) in the autistic group compared to the non-autistic groups (51%; p <.0001). All other differences between groups were non-significant. Based on data availability for the exposures, the sample sizes for models with each factor were as follows: autism diagnosis (n = 12,543), autism PGS (n = 7,283), social communication difficulties (n = 7,634), speech coherence (n = 7,860), sociability temperament (n = 9,723), and repetitive behaviour (n = 7,782).

**Table 2.** Demographic characteristics.

|  | Autistic | Non-Autistic | *p*-value |
| --- | --- | --- | --- |
| Total *n* | 150 | 13,886 | - |
| Sex *n* (%) <sup>a</sup> |  |  |  |
| Female | 32 (21) | 6825 (49) | <.0001 |
| Male |  |  |  |
| Ethnicity *n* <sup>a</sup> |  |  |  |
| Minoritised ethnicity | 5 | 595 | .56 |
| White | 133 | 11,136 |  |
| Maternal EPDS score, mean (SD) <sup>b</sup> | 6.25 (5.04) | 6.07 (4.80) | .70 |
| Maternal CCEI anxiety score, mean (SD) <sup>b</sup> | 3.64 (3.42) | 3.43 (3.33) | .49 |
| Financial difficulties score, mean (SD) <sup>b</sup> | 2.90 (3.46) | 2.92 (3.54) | .93 |
| Maternal highest educational attainment *n* <sup>a</sup> |  |  |  |
| CSE/None | 26 | 2415 | .53 |
| Vocational | 8 | 1191 |  |
| O Level | 50 | 4117 |  |
| A Level | 33 | 2658 |  |
| Degree | 21 | 1532 |  |
| Maternal social class *n* <sup>a</sup> |  |  |  |
| Professional | 7 | 562 | .38 |
| Managerial or technical | 43 | 3031 |  |
| Skilled (non-manual) | 46 | 4127 |  |
| Partly skilled, unskilled, skilled manual | 17 | 1928 |  |

<sup>a</sup> Pearson's chi-squared test for homogeneity, <sup>b</sup> Independent samples t-test.
EDPS: Edinburgh Postnatal Depression Scale; CCEI: Crown-Crisp Experiential Index.
**Note.** Sample sizes vary for each characteristic depending on data availability.

**Table 3.** Associations of key factors related to autism with night-time sleep duration trajectories.

| Factors | Shorter sleep duration vs<br>Intermediate-longer sleep duration |  | Longer sleep duration vs<br>Intermediate-longer sleep duration |  | Intermediate-shorter sleep duration<br>vs Intermediate-longer sleep duration |  |
| --- | --- | --- | --- | --- | --- | --- |
|  | OR (95% CI) | p-value | OR (95% CI) | p-value | OR (95% CI) | p-value |
| Autism diagnosis | 6.64 (3.40 – 12.98) | < **0.0001** | 0.44 (0.10 – 1.87) | 0.27 | 1.89 (1.02 – 3.52) | **0.04** |
| Autism polygenic score | 1.02 (0.90 – 1.17) | 0.74 | 0.99 (0.90 – 1.09) | 0.81 | 1.04 (0.96 – 1.14) | 0.33 |
| Social communication difficulties | 1.10 (1.06 – 1.13) | < **0.0001** | 0.99 (0.95 – 1.02) | 0.52 | 1.02 (1.00 – 1.05) | **0.05** |
| High social communication difficulties <sup>a</sup> | 2.55 (1.74 – 3.75) | < **0.0001** | 1.18 (0.88 – 1.60) | 0.27 | 1.04 (0.71 – 1.53) | 0.86 |
| Repetitive behaviour | 1.34 (1.11 – 1.61) | **0.002** | 0.86 (0.72 – 1.03) | 0.10 | 1.00 (0.89 – 1.13) | 0.97 |
| High repetitive behaviour <sup>a</sup> | 1.81 (1.03 – 3.21) | **0.04** | 0.84 (0.57 – 1.26) | 0.41 | 0.88 (0.55 – 1.41) | 0.60 |
| Speech coherence | 0.86 (0.79 – 0.94) | **0.001** | 1.01 (0.95 – 1.06) | 0.86 | 1.02 (0.97 – 1.07) | 0.51 |
| Low speech coherence <sup>a</sup> | 2.28 (1.17 – 4.44) | **0.02** | 0.91 (0.62 – 1.34) | 0.64 | 0.85 (0.61 – 1.17) | 0.31 |
| Sociability temperament | 1.03 (0.98 – 1.08) | 0.26 | 1.02 (0.99 – 1.05) | 0.25 | 1.01 (0.99 – 1.04) | 0.24 |
| Low sociability <sup>a</sup> | 1.09 (0.74 – 1.61) | 0.67 | 0.88 (0.65 – 1.19) | 0.40 | 0.98 (0.78 – 1.24) | 0.89 |

<sup>a</sup> Dichotomised autistic trait variables. OR; Odds Ratio.
**Note.** All models were adjusted for covariates except the autism PGS model which was not adjusted. Analysis was adjusted for the following confounders: sex, maternal postnatal depression, maternal anxiety, maternal occupational class, mother's highest level of education, financial problems, and epilepsy diagnosis.

Pearson’s correlation analyses revealed that social communication difficulties were weak-to-moderately correlated with speech coherence (r =-0.40, *p* < 0.001), and repetitive behaviour (r = 0.25, *p* < 0.001). Speech coherence and repetitive behaviour had a similar size correlation (r =-0.30, *p* < 0.001). Correlations between other trait combinations were either non-significant or very weak (r < 0.1). Full correlation results can be found in Table S3 of the supplement.

### Night-time sleep duration trajectories

Fit indices for all models assessed (2-5 classes) are presented in Table S4 in the supplement. BIC value decreased each time a class was added, which is commonly observed in large samples (Wiggins et al., 2015). Entropy was highest for the five-class model, but differences between the four-and five-class solutions were minimal, indicating similar classification accuracy. The VLMR was significant for the two-, three-, and four-class models (all *p* ≤ 0.01) but not for the five-class model. Taken together, these indices supported the four-class model as the best-fitting solution, which was therefore selected as the optimal model after confirming adequate class sizes (>2%). Trajectories for all tested models are presented in the supplement (Figure S1).

The latent classes of trajectories from the final 4-class model are presented in Figure 1. All trajectories displayed a persistently decreasing trend from childhood to adolescence. The following trajectories were derived: 1. Shorter sleep duration (n = 512 [4.1%]), 2. Longer sleep duration (n = 1654 [13.1%]), 3. Intermediate-shorter sleep duration (n = 3630 [28.8%]), and 4. Intermediate-longer sleep duration (n = 6825 [54.1%]). The intermediate-longer sleep duration trajectory was used as the reference class when examining the associations between sleep trajectories and autism related factors as it had the largest class membership.

**Figure 1.**
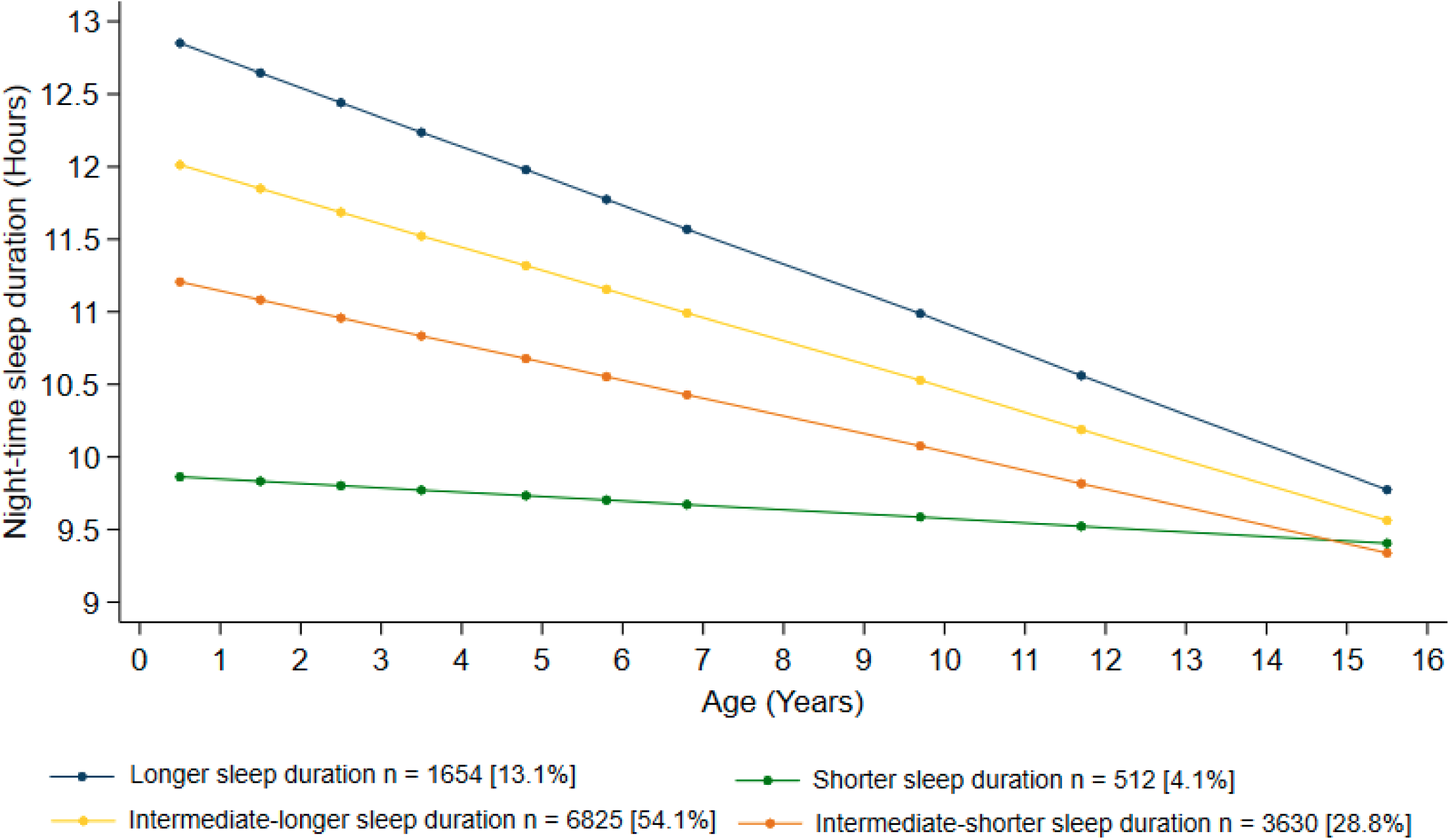
Night-time sleep duration trajectories from a 4-class solution.

### Associations between exposures and night-time sleep duration trajectories

Results of the adjusted multinomial logistic regression are presented for each sleep trajectory; full results are displayed in Table 2. Results from the unadjusted models were largely aligned with adjusted model results and are presented in the supplementary material (Table S5).

### Shorter sleep duration trajectory

Having an autism diagnosis was associated with increased likelihood of having a shorter sleep duration trajectory (OR = 6.64, 95%CI [3.40 – 12.98], p < 0.0001) compared to an intermediate-longer sleep duration trajectory (reference class). Among the autistic traits, being in the highest scoring group for social communication difficulties (OR = 2.55, 95% CI [1.74–3.75], *p* < 0.0001) and repetitive behaviour (OR = 1.81, 95% CI [1.03–3.21], *p* = 0.04), as well as being in the lowest scoring group for speech coherence (OR = 2.28, 95% CI [1.17–4.44], *p* = 0.02), were associated with increased odds of having a shorter sleep duration trajectory relative to the intermediate-longer trajectory (reference class). Continuous (total) scores for these three traits showed associations in the same direction. In contrast, being in the lowest scoring group for sociability temperament was not associated with a shorter sleep duration trajectory (OR = 1.09, 95%CI [0.74 – 1.61], p = 0.67), nor was the total score for this trait (OR = 1.03, 95%CI [0.98 – 1.08], p = 0.26). Lastly, the autism PGS was not significantly associated with a shorter sleep duration trajectory relative to an intermediate-longer trajectory (OR = 1.02, 95%CI [0.90– 1.17], p = 0.74).

### Intermediate-shorter sleep duration trajectory

Having an autism diagnosis was associated with increased likelihood of having an intermediate-shorter sleep duration trajectory (OR = 1.89, 95%CI [1.02 – 3.52], p = 0.04) relative to an intermediate-longer trajectory (reference class). None of the autistic traits, when examined as binary trait indicators, were associated with an intermediate-shorter sleep duration trajectory. However, higher total scores for social communication difficulties were marginally associated with increased odds of having an intermediate-shorter trajectory relative to the reference class (OR = 1.89, 95% CI [1.02–3.52], *p* = 0.04). No other total trait scores, nor autism PGS, were associated with this trajectory.

### Longer sleep duration trajectory

Autism diagnosis, autistic traits, and autism PGS were not associated with a longer sleep duration trajectory class relative to the intermediate-longer trajectory (reference class).

## Discussion

We identified four distinct night-time sleep duration trajectories within a general population cohort that included autistic individuals. We further examined whether an autism diagnosis, autistic traits, and autism PGS were associated with these sleep trajectories. We found that an autism diagnosis was associated with an increased likelihood of following sleep duration trajectories indicative of poorer sleep patterns – specifically shorter and intermediate-shorter sleep duration – relative to the intermediate-longer trajectory (the reference class). Among the autistic traits examined, higher levels of repetitive behaviour, lower speech coherence, and greater social communication difficulties were also associated with an increased likelihood of having a shorter sleep duration trajectory compared to an intermediate-longer trajectory. In contrast, sociability and autism PGS were not associated with any of the identified night-time sleep duration trajectories relative to the reference class.

The findings from this study were consistent with two previous studies in ALSPAC that also used LCGA to identify four distinct subclasses of night-time sleep duration trajectories with similar overall trajectory shapes and comparable proportions of participants in each class (Manitsa et al., 2024; Morales-Muñoz et al., 2024). In the current study, differences between night-time sleep duration trajectories were most pronounced in early childhood and gradually attenuated with age, resulting in more similar sleep durations across trajectories by 16 years. One possible explanation is that night-time sleep duration at the final assessment (15.5 years) was self-reported, whereas earlier assessments were parent-reported, which might result in measurement error discrepancies between the earlier time points and the final one. However, a previous ALSPAC study found that excluding this final time point yielded similar results (Manitsa et al., 2024). A recent systematic review of research on group-based sleep trajectories reported that this pattern of diminishing differences between trajectories with age is common across studies (Wang et al., 2024). This could be a reflection of greater initial variability in circadian rhythm development in younger children which stabilises with age, and external factors such as school that enforce more consistent sleep routines as children grow older.

As hypothesised, we found that having an autism diagnosis was associated with an increase in the likelihood of having a shorter sleep duration trajectory compared to an intermediate-longer trajectory, which adds to a growing body of evidence demonstrating that autistic individuals are more likely to experience long-term patterns of sleep disturbance (Baker et al., 2023; Jovevska et al., 2020; Verhoeff et al., 2018). Additionally, specific autistic traits were associated with a shorter sleep duration trajectory. These traits showed only weak intercorrelations suggesting that they may exert independent influences on sleep patterns. Further research utilising repeated measures of autistic traits and sleep is required to establish any causal links. Nonetheless, we speculate on several potential mechanistic implications below.

Much of the existing research linking autistic traits to sleep problems has been cross-sectional (Whelan et al., 2024), limiting insight into how these associations unfold across development. Here, we found that three parent-reported autistic traits – greater social communication difficulties, lower speech coherence, and higher levels of repetitive behaviour – were associated with an increased likelihood of following a persistently shorter sleep duration trajectory (compared to an intermediate-longer trajectory), which remained lower than other trajectories from infancy through approximately 12 years of age. These findings suggest that mechanisms linking these autistic traits to sleep problems may operate differently across developmental stages. While current theories aiming to explain autistic people’s sleep problems often emphasise childhood-specific factors such as bedtime routines (Mazzone et al., 2018), our results highlight the importance of considering age-scalable mechanisms that extend beyond periods when sleep is largely regulated by parental structure. For example, pragmatic language difficulties, captured here as low speech coherence, could influence sleep not only by affecting comprehension of bedtime instructions in childhood, but also by limiting the expression of emotional distress, thereby interfering with relaxation and sleep onset later in development.

Similarly, behavioural profiles characterised by repetitive motor behaviours (as measured in this study) may be accompanied by repetitive cognitive processes, such as intrusive thoughts or rumination at bedtime, which could continue to disrupt sleep across development (Harvey et al., 2005). Insistence on elaborate routines at bedtime is another element of the repetitive behaviour domain which may directly delay sleep onset and therefore reduce sleep duration. Evidence thus far suggests that the association between repetitive behaviours and sleep is complex and behaviour-specific. A recent systematic review reported stronger associations between sleep problems and certain repetitive behaviours, such as ritualistic behaviours, than with others, such as self-injurious behaviours (Passarini et al., 2025). Anxiety has also been proposed as a potential mediating factor, with repetitive behaviours and routines reflecting a preference for sameness or ritualism possibly serving to regulate anxiety, although evidence remains inconclusive (MacDuffie et al., 2020). Therefore, clarifying the role of different forms of repetitive behaviour in this context may be helpful for developing behavioural sleep interventions for autistic individuals.

While both the presence of parent-reported social communication difficulties and low sociability capture social characteristics associated with autism, only social communication difficulties were associated with a shorter sleep duration trajectory. Although sociability was measured at age 3 years, this trait has been shown to be stable over time in the ALSPAC cohort (Bould et al., 2013), making the early timing of assessment unlikely to explain this null finding. A key distinction between these constructs is that social communication difficulties reflect challenges with social reciprocity (in a majority non-autistic society) and interpreting normative social cues, whereas low sociability captures a preference for solitary activities. Therefore, a lack of relation between sociability and sleep calls somewhat into question the hypothesis that missing the shared activities that help align circadian rhythms (e.g. meals, sports) is a key explanation for autistic people’s sleep disruption (Deliens & Peigneux, 2019; Geoffray et al., 2016). One interpretation is that a preference for solitude is not inherently distressing for autistic individuals and therefore may not be disruptive to sleep (Calder et al., 2013), while the daily experience of challenges communicating and interacting with others may disrupt sleep. As in the case of repetitive behaviours, this link could be mediated by anxiety, where sleep is disrupted by anxieties stemming from challenging social interactions experienced during the completed day, or anticipating the social demands of tomorrow.

Finally, we found no evidence that autism PGS was associated with shorter or longer night-time sleep duration trajectories relative to the intermediate-longer trajectory. This is consistent with previous studies that reported no associations between genetic liability for autism and cross-sectional measures of sleep disturbance or clinical insomnia diagnoses (Niarchou et al., 2022; Ohi et al., 2021). One possible explanation is that current polygenic scores capture only a modest proportion of the genetic liability for autism (approximately 12%) (Grove et al., 2019), and future genome-wide association studies with greater power may better elucidate genetic contributions to sleep outcomes. Although existing evidence suggests that autism PGS is not a strong predictor of sleep duration, a recent study reported an association between autism PGS and evening chronotype (Fahey & Lopez, 2024). This raises the possibility that while genetic liability for autism can influence circadian preference, the behavioural manifestation of this might be moderated by environmental factors or behavioural strategies that compensate to preserve sleep duration, or that such genetic effects may be more relevant to other aspects of sleep. Together, these findings highlight the need for further research to clarify how genetic risk for autism interacts with environmental and biological factors to shape sleep patterns across development.

### Strengths and Limitations

Key strengths of this study include the large sample size, the availability of prospectively collected repeated measures of sleep and the population-based design of the study – which helps minimise the risk of recall and selection bias. Furthermore, we took an inclusive approach to delineate our autistic sample by using multiple, independent sources, to help address the limitation that autistic individuals (particularly those diagnosed later in life) are likely to be underrepresented in this cohort. In addition, rich covariable information were available in the dataset, which allowed us to account for multiple confounding factors that could impact the relationship between autism and sleep.

When interpreting findings, however, a crucial limitation is that measuring these traits in a general population sample might not adequately capture their manifestations and associated behaviours in the autistic population (Sasson & Bottema-Beutel, 2022). As such, whilst these findings offer preliminary insights into potential mechanisms underpinning sleep problems in autistic individuals, the specific implications of the findings for the autistic population are limited.

The reliance on parent-and self-reported sleep measures is also an important limitation, as discrepancies between subjective and objective sleep estimates have been reported, including both over-and underestimation (O’Sullivan et al., 2023). Such inaccuracies may increase with age, particularly during adolescence when parental oversight decreases. Thus, future research in this area would benefit from tracking sleep over time using objective methods (e.g., actigraphy or radar technology) as well as subjective assessments. In addition, sleep duration from 6 months to 11 years was derived from estimated bedtimes and wake times, which do not capture sleep onset latency or wakefulness after sleep onset – both commonly reported sleep difficulties in autistic individuals (Richdale & Schreck, 2009).

A further limitation of this study is that elevated autistic traits in a general population sample may partially act as a proxy for autism diagnosis. As a result, associations observed between autistic traits and long-term sleep trajectories may reflect the influence of other unmeasured autism-related factors, such as sensory sensitivities, anxiety, or gastrointestinal difficulties, which could represent more proximal mechanisms linking autism and sleep problems. Replication of these findings in an autism-specific sample, with adjustment for such co-occurring factors, may help to clarify these associations.

## Conclusion

By identifying heterogeneous night-time sleep duration trajectories and linking shorter sleep duration trajectories to autism diagnosis and specific autism-related traits, our findings suggest that multiple, trait-specific mechanisms may contribute to long-term sleep difficulties for autistic individuals. These results emphasise the importance of considering developmental scalability and individual differences when investigating sleep problems in autism.

## Data Availability

All data from the current study is available for bona fide researchers here: https://www.bristol.ac.uk/alspac/researchers/our-data/

## Notes

**Funding** The UK Medical Research Council, Wellcome (MR/Z505924/1), and the University of Bristol provide core support for ALSPAC. This study was funded by the Wellcome Trust (218493/Z/19/Z; 227063/Z/23/Z). R.Z. and A.S.F.K are the guarantors of this work, and S.M. and A.S.F.K are supported by a Wellcome Early Career Award (227063/Z/23/Z). Genome-wide genotyping was conducted by the Wellcome Sanger Institute and LabCorp with support from 23andMe. For Open Access, a CC BY licence applies to the Author Accepted Manuscript. A full list of ALSPAC funding is available on the ALSPAC website. (http://www.bristol.ac.uk/alspac/external/documents/grant-acknowledgements.pdf).

### Competing Interest Statement

The authors have declared no competing interest.

### Funding Statement

The UK Medical Research Council, Wellcome (MR/Z505924/1), and the University of Bristol provide core support for ALSPAC. This study was funded by the Wellcome Trust (218493/Z/19/Z; 227063/Z/23/Z). R.Z. and A.S.F.K are the guarantors of this work, and S.M. and A.S.F.K are supported by a Wellcome Early Career Award (227063/Z/23/Z). Genome-wide genotyping was conducted by the Wellcome Sanger Institute and LabCorp with support from 23andMe. For Open Access, a CC BY licence applies to the Author Accepted Manuscript. A full list of ALSPAC funding is available on the ALSPAC website. (http://www.bristol.ac.uk/alspac/external/documents/grant-acknowledgements.pdf).

### Author Declarations

The Avon Longitudinal Study of Parents and Children (ALSPAC) Law and Ethics committee and the NHS Haydock Research Ethics Committee gave ethical approval for this study (Approval Reference Number: 10/H1010/70).

