## Supplementary Materials for "Associations of autism diagnosis, traits, and genetic liability with subsequent night-time sleep duration trajectories from infancy to adolescence"

Supplementary Material

### S1. Genotyping Information

ALSPAC children were genotyped using the Illumina HumanHap550 quad chip genotyping platforms by 23andme subcontracting the Wellcome Trust Sanger Institute, Cambridge, UK and the Laboratory Corporation of America, Burlington, NC, US. The resulting raw genome-wide data were subjected to standard quality control methods on 9,915 subjects and 550,000 SNPs. Individuals were excluded on the basis of gender mismatches; minimal or excessive heterozygosity; disproportionate levels of individual missingness (>3%) and insufficient sample replication (IBD < 0.8). Population stratification was assessed by multidimensional scaling analysis and compared with Hapmap II (release 22) European descent (CEU), Han Chinese, Japanese and Yoruba reference populations; all individuals with non-European ancestry were removed by removing samples that clustered outside the CEU HapMap2 population using this multidimensional scaling of genome-wide IBS pairwise distances. SNPs with a minor allele frequency of < 1%, a call rate of < 95% or evidence for violations of Hardy-Weinberg equilibrium (P < 5E-7) were removed. Cryptic relatedness was measured as proportion of identity by descent (IBD > 0.1). Related subjects that passed all other quality control thresholds were retained during subsequent phasing and imputation. 9,115 subjects and 500,527 SNPs passed these quality control filters.

ALSPAC mothers were genotyped using the Illumina human660W-quad array at Centre National de Genotypage (CNG) and genotypes were called with Illumina GenomeStudio. PLINK (v1.07) was used to carry out quality control measures on an initial set of 10,015 subjects and 557,124 directly genotyped SNPs. SNPs were removed if they displayed more than 5% missingness or a Hardy-Weinberg equilibrium P value of less than 1.0e-06. Additionally, SNPs with a minor allele frequency of less than 1% were removed. Samples were excluded if they displayed more than 5% missingness, had indeterminate X chromosome heterozygosity or extreme autosomal heterozygosity. Samples showing evidence of population stratification were identified by multidimensional scaling of genome-wide identity by state pairwise distances using the four HapMap populations as a reference, and then excluded. Cryptic relatedness was assessed using a IBD estimate of more than 0.125 which is expected to correspond to roughly 12.5% alleles shared IBD or a relatedness at the first cousin level. Related subjects that passed all other quality control thresholds were retained during subsequent phasing and imputation. 9,048 subjects and 526,688 SNPs passed these quality control filters.

We combined 477,482 SNP genotypes in common between the sample of mothers and sample of children. We removed SNPs with genotype missingness above 1% due to poor quality (11,396 SNPs removed) and removed a further 321 subjects due to potential ID mismatches. This resulted in a dataset of 17,842 subjects containing 6,305 duos and 465,740 SNPs (112 were removed during liftover and 234 were out of HWE after combination). We estimated haplotypes using ShapeIT (v2.r644) which utilises relatedness during phasing. We obtained a phased version of the 1000 genomes reference panel (Phase 1, Version 3) from the Impute2 reference data repository (phased using ShapeIT v2.r644, haplotype release date Dec 2013). Imputation of the target data was performed using Impute V2.2.2 against the reference panel (all polymorphic SNPs excluding singletons), using all 2186 reference haplotypes (including non-Europeans). This resulted in 28,699,419 SNPs, with 8,282,911 SNPs with a MAF >0.01 and info score of >0.8.

This gave 8,237 eligible children and 8,196 eligible mothers with available genotype data after exclusion of related subjects using cryptic relatedness measures described previously. Only ALSPAC children were used in the following analyses.

**Table S1.** Covariate measurement.

| Covariate | Source |
|---|---|
| Sex | ALSPAC study administrative variable |
| Maternal postnatal depression | Edinburgh Postnatal Depression Scale administered at 8 weeks (completed by mother) |
| Maternal anxiety | Crisp Crown Experiential Index Anxiety subscale II administered at 8 weeks (completed by mother) |
| Epilepsy diagnosis | ALSPAC questionnaires administered at 1.5, 2.5, 3.5, 5, 6, 7, 9, and 11 years asked parents whether their child had ever experienced a seizure due to epilepsy. Participants were classified as having epilepsy if a parent reported ‘Yes’ at any assessment |
| Maternal occupational class | ALSPAC questionnaire administered at 32 weeks’ gestation (completed by the mother) asked respondents to indicate the category that best described their current occupation: professional; managerial or technical; skilled (non-manual); partly skilled; skilled manual; or unskilled. Due to small cell sizes, the latter three categories were combined for analysis |
| Highest level of maternal educational attainment | ALSPAC questionnaire administered at 32 weeks’ gestation (completed by the mother) asked respondents to indicate the category that best described their highest level of education: CSE/none; vocational; O level; A level; or degree. |
| Financial problems | ALSPAC questionnaire administered at 32 weeks’ gestation (completed by the mother) asked questions about financial difficulties in the household. ALSPAC derived a financial difficulties score (ranging 0-15) based on the responses. |

**Table S2.** Sensitivity analysis results showing associations of key factors related to autism with night-time sleep duration trajectories in the subsample with data available for all factors (n = 3844).

| Factors | Shorter sleep duration vs Intermediate-longer sleep duration | | Longer sleep duration vs Intermediate-longer sleep duration | | Intermediate-shorter sleep duration vs Intermediate-longer sleep duration | |
|---|---|---|---|---|---|---|
| | **OR (95% CI)** | **p-value** | **OR (95% CI)** | **p-value** | **OR (95% CI)** | **p-value** |
| Autism diagnosis | 3.30 (1.32 – 8.31) | **0.01** | 0.30 (0.04 – 2.03) | 0.22 | 1.29 (0.57 – 2.92) | 0.55 |
| Autism polygenic score | 1.13 (0.95 – 1.34) | 0.16 | 0.96 (0.85 – 1.09) | 0.52 | 1.08 (0.98 – 1.19) | 0.11 |
| Social communication difficulties | 1.13 (1.09 – 1.17) | **< 0.0001** | 0.97 (0.92 – 1.03) | 0.36 | 1.04 (1.00 – 1.08) | 0.09 |
| High social communication difficulties [a] | 3.57 (2.26 – 5.63) | **< 0.0001** | 1.06 (0.65 – 1.73) | 0.82 | 1.39 (0.88 – 2.21) | 0.16 |
| Repetitive behaviour | 1.47 (1.14 – 1.89) | **0.003** | 0.77 (0.61 – 0.98) | 0.03 | 0.98 (0.85 – 1.15) | 0.83 |
| High repetitive behaviour [a] | 1.76 (0.77 – 4.04) | 0.18 | 0.48 (0.20 – 1.16) | 0.10 | 0.73 (0.46 – 1.17) | 0.19 |
| Speech coherence | 0.89 (0.81 – 0.98) | **0.02** | 1.04 (0.95 – 1.14) | 0.43 | 1.01 (0.95 – 1.08) | 0.75 |
| Low speech coherence [a] | 1.82 (1.03 – 3.24) | **0.04** | 0.74 (0.37 – 1.47) | 0.39 | 0.91 (0.59 – 1.40) | 0.66 |
| Sociability temperament | 0.98 (0.89 – 1.08) | 0.70 | 1.04 (0.99 – 1.08) | 0.09 | 1.02 (0.98 – 1.06) | 0.37 |
| Low sociability [a] | 1.70 (1.06 – 2.73) | 0.04 | 0.84 (0.55 – 1.29) | 0.43 | 0.90 (0.64 – 1.27) | 0.56 |

[a] Dichotomised autistic trait variables. OR; Odds Ratio

**Table S3.** Sensitivity analysis results showing associations of key factors related to autism with night-time sleep duration trajectories in the subsample with data available for all factors (n = 3844).

| Factors | Mean age (Years) | Mean Score (SD) | Total n included in analyses | Social communication difficulties | Speech coherence | Sociability temperament | Repetitive Behaviour |
|---|---|---|---|---|---|---|---|
| Autism diagnosis | - | - | 12,543 | - | - | - | - |
| Autism polygenic score | - | - | 7,283 | - | - | - | - |
| Social communication difficulties | 7.66 | 2.80 (3.73) | 7,634 | - | - | - | - |
| Speech coherence | 9.65 | 34.81 (2.07) | 7,860 | -0.40** | - | - | - |
| Sociability temperament | 3.21 | 18.21 (3.11) | 9,723 | -0.002 | 0.087** | - | - |
| Repetitive behaviour | 5.79 | 4.34 (0.65) | 7,782 | 0.25** | -0.30** | -0.042* | - |

**Table S4.** Model fit indices for 2-5 class solutions for night-time sleep duration trajectories

| Night-time sleep duration models | No. of participants assigned to class *n* (%) | BIC | Entropy | VLMR-*p* |
|---|---|---|---|---|
| 2 classes | Class 1 – 4789 (37.9)<br>Class 2 – 7831 (62.1) | 221281 | 0.609 | <0.00001 |
| 3 classes | Class 1 – 1544 (12.2)<br>Class 2 – 3811 (30.2)<br>Class 3 – 7265 (57.6) | 218500.2 | 0.637 | <0.00001 |
| **4 classes** | **Class 1 – 512 (4.1)**<br>**Class 2 – 1654 (13.1)**<br>**Class 3 – 3630 (28.8)**<br>**Class 4 – 6825 (54.1)** | **217530.9** | **0.65** | **0.0081** |
| 5 classes | Class 1 – 360 (2.9)<br>Class 2 – 462 (3.7)<br>Class 3 – 1658 (13.1)<br>Class 4 – 3374 (26.7)<br>Class 5 – 6766 (53.6) | 217064 | 0.676 | 0.1425 |

BIC; Bayesian information criterion, VLMR; Vuong-Lo-Mendell-Rubin likelihood ratio test.

**Table S5.** Unadjusted associations of key factors related to autism with night-time sleep trajectories.

| Factors | Shorter sleep duration vs Intermediate-longer sleep duration | | Longer sleep duration vs Intermediate-longer sleep duration | | Intermediate-shorter sleep duration vs Intermediate-longer sleep duration | |
|---|---|---|---|---|---|---|
| | **OR (95% CI)** | **p-value** | **OR (95% CI)** | **p-value** | **OR (95% CI)** | **p-value** |
| Autism diagnosis | 6.27 (3.55 – 11.09) | < **0.0001** | 0.47 (0.14 – 1.57) | 0.22 | 2.00 (1.15 – 3.49) | **0.01** |
| Autism polygenic score | 1.02 (0.90 – 1.17) | 0.74 | 0.99 (0.90 – 1.09) | 0.81 | 1.04 (0.96 – 1.14) | 0.33 |
| Social communication difficulties | 1.12 (1.06 – 1.15) | < **0.0001** | 0.99 (0.95 – 1.02) | 0.39 | 1.04 (1.02 – 1.06) | < **0.0001** |
| High social communication difficulties [a] | 3.05 (2.15 – 4.32) | < **0.0001** | 1.41 (1.09 – 1.82) | **0.008** | 1.13 (0.81 – 1.58) | 0.46 |
| Repetitive behaviour | 1.51 (1.28 – 1.77) | < **0.0001** | 0.92 (0.79 – 1.07) | 0.26 | 1.01 (0.91 – 1.11) | 0.93 |
| High repetitive behaviour [a] | 2.66 (1.55 – 4.56) | < **0.0001** | 1.12 (0.81 – 1.55) | 0.50 | 0.16 (0.79 – 1.72) | 0.45 |
| Speech coherence | 0.85 (0.80 – 0.90) | < **0.0001** | 1.00 (0.95 – 1.04) | 0.83 | 1.00 (0.96 – 1.04) | 0.99 |
| Low speech coherence [a] | 2.49 (1.58 – 3.92) | < **0.0001** | 0.96 (0.69 – 1.34) | 0.82 | 0.90 (0.64 – 1.26) | 0.54 |
| Sociability temperament | 1.01 (0.97 – 1.06) | 0.62 | 1.01 (0.98 – 1.04) | 0.54 | 1.02 (1.00 – 1.05) | **0.02** |
| Low sociability [a] | 1.15 (0.81 – 1.63) | 0.42 | 1.06 (0.82 – 1.37) | 0.66 | 0.97 (0.79 – 1.18) | 0.73 |

[a] Dichotomised autistic trait variables. OR; Odds Ratio

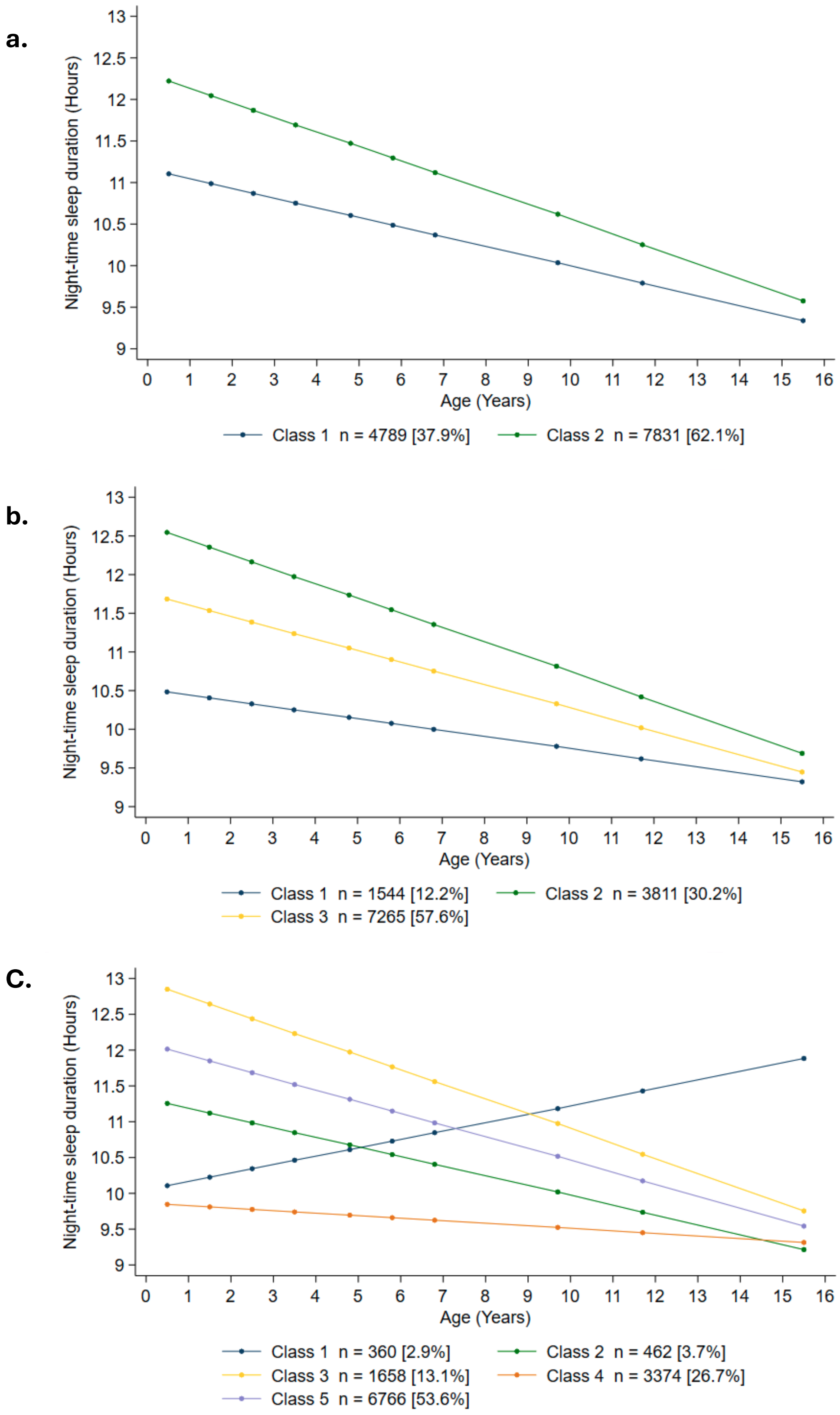


**Figure S1.** Trajectories of night-time sleep duration from models with **a.** 2 class, **b.** 3 class, and **c.** 5 class solutions.
